# AERO: An AI Agent for Adaptive Eligibility Refinement and Optimization of Clinical Trial Criteria in Real-World Trial Emulation

**DOI:** 10.64898/2026.04.30.26352142

**Authors:** Xiaodi Li, Jose James, Patricia Pellikka, Nansu Zong

**Affiliations:** Department of Artificial Intelligence and Informatics, Mayo Clinic, Rochester, MN, USA; Mayo Clinic School of Graduate Medical Education, Rochester, MN, USA; Department of Cardiovascular Medicine, Mayo Clinic, Rochester, MN, USA

## Abstract

Randomized controlled trials (RCTs) provide high internal validity but often rely on restrictive eligibility criteria that limit generalizability and complicate real-world trial emulation. We propose AERO (AI Agent for Adaptive Eligibility Refinement and Optimization), an agentic framework that systematically adapts clinical trial eligibility criteria for application to electronic health record data. AERO integrates external clinical knowledge sources and large language model–based reasoning to classify criteria as strict inclusion, safety exclusion, confounder, or operational artifact. We evaluated AERO by emulating the WARCEF trial using Mayo Clinic Platform data restricted to the pre-trial completion period. Emulation with optimized criteria yielded a hazard ratio of 1.561 (p = 0.0605), consistent with the original neutral trial finding (HR = 1.01, p = 0.91). An ablation analysis demonstrated that eligibility handling decisions materially influence observed treatment effects. These results highlight the importance of systematic, knowledge-informed eligibility refinement in real-world evidence generation.

## Introduction

Randomized controlled trials (RCTs) are widely regarded as the gold standard for evaluating the efficacy and safety of medical interventions because randomization minimizes confounding and enables strong causal inference ^1, 2^. However, the design of trial protocols, including the specification of eligibility criteria, is typically manual and expert-driven, which makes systematic optimization and adaptation to real-world data challenging ^3, 4^. Consequently, treatment effects observed in trials may not reflect effectiveness in broader and more heterogeneous populations encountered in everyday care. Real-world evidence (RWE), derived from observational data sources such as electronic health records (EHRs), administrative claims, and clinical registries, complements RCTs by capturing outcomes in routine clinical settings and diverse patient populations ^5-7^. The increasing availability of large-scale healthcare data infrastructures has enabled RWE to play an expanding role in clinical research, health system evaluation, and regulatory decision-making ^5, 8-10^.

Target trial emulation has emerged as a principled framework for using observational data to approximate the design and causal estimand of a hypothetical randomized trial ^11-13^. By explicitly specifying eligibility criteria, treatment strategies, assignment procedures, follow-up periods, and outcome definitions, this approach aims to reduce biases inherent in naïve observational analyses. Nevertheless, a major obstacle to successful emulation lies in translating protocol-defined eligibility criteria into real-world data. Many trial criteria rely on measurements, assessments, or operational requirements that are unavailable, inconsistently recorded, or impractical within EHR systems. Applying these criteria verbatim can introduce substantial selection bias, reduce cohort size, or exclude clinically relevant patients, thereby distorting comparative effectiveness estimates ^14^.

Despite increasing interest in target trial emulation, adapting eligibility criteria for real-world data remains challenging. In practice, several limitations arise during the translation of protocol-defined eligibility criteria into EHR-based cohorts. The process of eligibility adaptation in trial emulation presents several disadvantages: (1) certain eligibility criteria that capture baseline risk or patient characteristics are often reinterpreted as confounders and adjusted for during statistical analysis rather than applied as strict filtering rules; (2) this reinterpretation process is typically performed manually by domain experts and relies heavily on subjective judgment; (3) researchers may relax restrictive eligibility criteria or reverse engineer trial populations within observational datasets to approximate the original trial cohort; (4) these adaptations frequently require manual decisions regarding which criteria should be relaxed, reclassified, or retained, making the process labor-intensive and difficult to reproduce; and (5) such ad hoc modifications may introduce domain expert bias or data-driven decisions that compromise the consistency and transparency of trial emulation.

To address these challenges, we propose AERO (AI Agent for Adaptive Eligibility Refinement and Optimization), an agentic framework designed to automate the adaptation of clinical trial eligibility criteria for real-world cohort construction. Built on an augmented large language model capable of multi-step reasoning and tool use, AERO integrates heterogeneous medical knowledge sources, including up-to-date clinical knowledge bases, medical language models, and structured drug information repositories, to systematically interpret and operationalize eligibility criteria in EHR environments. Specifically, AERO addresses the limitations of current trial emulation workflows by: (1) systematically identifying criteria that capture baseline risk and reclassifying them as confounders when appropriate rather than relying on subjective expert reinterpretation; (2) automating the evaluation of each eligibility criterion using protocol context, clinical knowledge, and data feasibility constraints to reduce reliance on manual expert judgment; (3) identifying overly restrictive criteria and recommending principled relaxation strategies without requiring ad hoc reverse engineering of trial populations; (4) standardizing the decision process for retaining, relaxing, reclassifying, or removing criteria, thereby improving reproducibility and scalability across studies; and (5) reducing domain expert bias and ad hoc data-driven adaptations through a transparent, knowledge-informed reasoning framework. Through this process, AERO produces eligibility criteria that preserve clinical intent while remaining operationally compatible with real-world EHR data, thereby improving the robustness, reproducibility, and validity of target trial emulation.

We evaluate AERO by emulating the WARCEF (Warfarin versus Aspirin in Reduced Cardiac Ejection Fraction) trial using real-world data from the Mayo Clinic Platform (MCP), a large multi-institutional EHR resource. The original WARCEF trial compared warfarin and aspirin in patients with heart failure and reduced ejection fraction (HFrEF) in sinus rhythm and found no statistically significant difference between treatments for the primary outcome ^15^. Using AERO-optimized eligibility criteria, we construct real-world cohorts from MCP data in the pre-trial completion period and estimate comparative effectiveness between treatments following target trial principles.

This work makes three principal contributions. (1) AERO represents, to our knowledge, one of the first AI agent– based approaches specifically designed for systematic refinement of clinical trial eligibility criteria for real-world trial emulation. (2) The framework explicitly integrates diverse external medical knowledge sources to support clinically grounded reasoning, rather than relying solely on data-driven filtering or heuristic rules. (3) We demonstrate a scalable and reproducible pipeline for translating RCT protocols into real-world studies using large EHR platforms. Together, these contributions advance the methodological foundation of AI-enabled evidence generation and illustrate how agentic systems can help bridge the gap between controlled clinical trials and real-world clinical practice.

## Methods

### Overview

We developed AERO, an agentic framework that automatically adapts clinical trial eligibility criteria for real-world trial emulation. The pipeline consists of five sequential stages: (1) extraction of original eligibility criteria, (2) retrieval of external medical knowledge, (3) AI-driven criteria optimization, (4) real-world cohort construction and trial emulation, and (5) outcome analysis (Figure 1).

**Figure 1.**
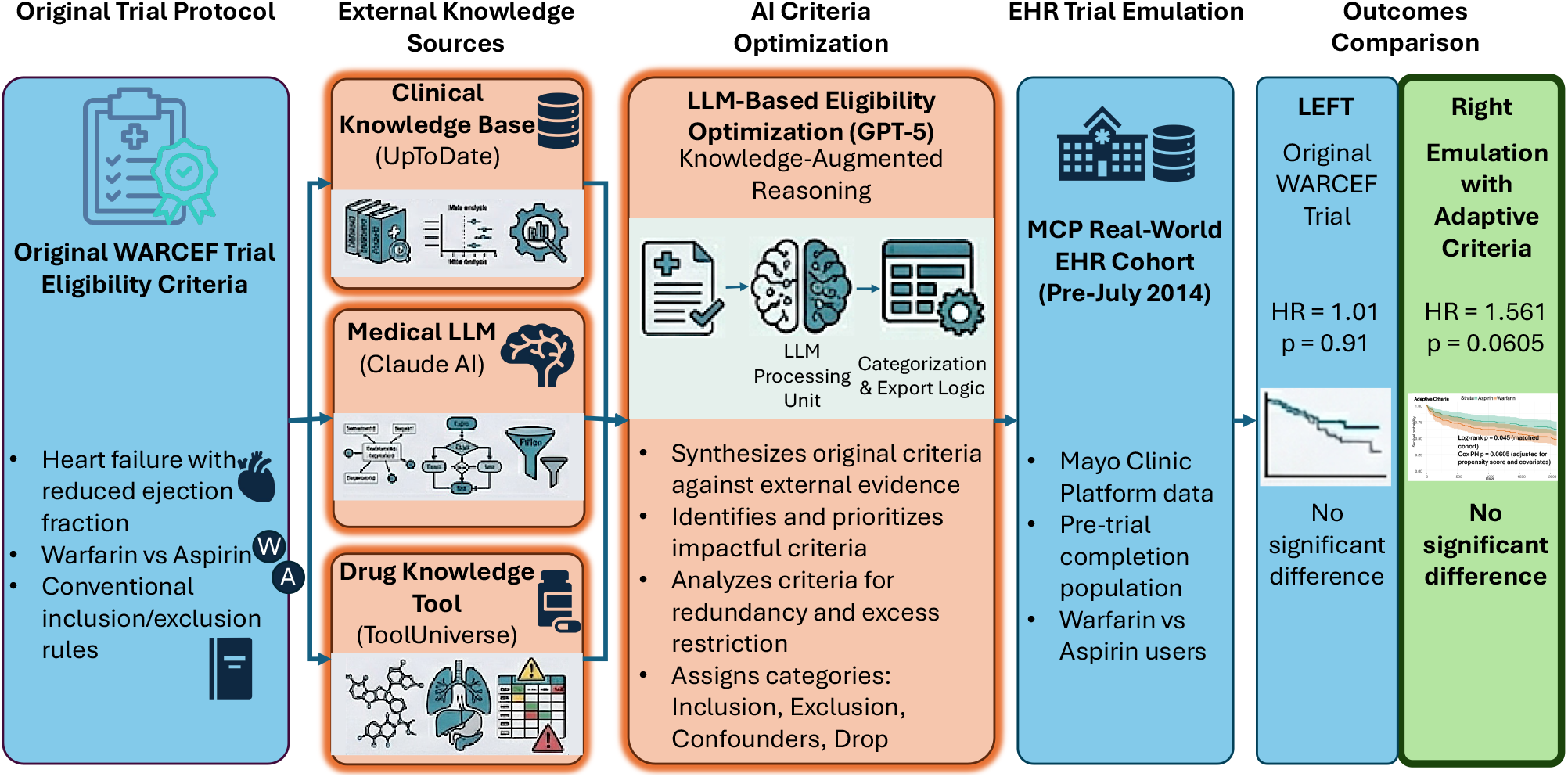
Overview of the AERO Framework for Adaptive Eligibility Criteria Optimization and Trial Emulation. The AERO pipeline begins with extraction of the original eligibility criteria from the WARCEF trial protocol. External clinical knowledge is then retrieved from multiple sources, including a clinical knowledge base (UpToDate ^16^), a medical large language model (Claude AI ^17^), and a drug knowledge tool (ToolUniverse ^18^). These knowledge sources, together with the original criteria, are provided to a knowledge-augmented language model (GPT-5 ^19^), which analyzes each criterion and categorizes it as inclusion, exclusion, confounder, or drop based on clinical relevance, feasibility in EHR data, and potential for selection bias. The optimized criteria are subsequently applied to real-world data from the MCP to construct treatment cohorts for target trial emulation in the pre-trial completion period. Comparative effectiveness between warfarin and aspirin is evaluated using survival analysis. The results show that emulation using adaptive criteria (HR = 1.561, p = 0.0605) yields conclusions consistent with the original WARCEF trial (HR = 1.01, p = 0.91), demonstrating that AERO preserves the trial’s overall finding of no significant difference while improving real-world applicability.

### Extraction of Original Eligibility Criteria

The original eligibility criteria were obtained directly from the publicly available trial registration on ClinicalTrials.gov. Inclusion and exclusion criteria were copied from the trial protocol description, preserving their original wording to maintain fidelity to the source design. These criteria serve as the baseline specification of the target trial population.

### Retrieval of External Knowledge Sources

To support clinically informed reasoning, AERO incorporates heterogeneous external knowledge sources relevant to the trial context. Knowledge retrieval was performed based on the trial description, disease area, and interventions.

Three complementary sources were used: (1) Clinical knowledge base: Up-to-date clinical information retrieved from UpToDate ^16^, providing disease background, treatment indications, contraindications, and standard-of-care considerations. (2) Medical large language model: Contextual clinical interpretation generated using Claude AI ^17^. (3) Drug knowledge tool: Structured pharmacological and safety information obtained from ToolUniverse ^18^.

These sources collectively provide domain knowledge regarding patient characteristics, treatment mechanisms, contraindications, and real-world clinical practice patterns.

### AI-Driven Eligibility Criteria Optimization

The original criteria and retrieved knowledge were jointly provided as input to a large language model (GPT-5 ^19^) operating as the reasoning core of AERO. The model analyzes each criterion and assigns it to one of four categories: (1) Inclusion: Essential characteristics defining the target population. (2) Exclusion: Safety-related or clinically necessary restrictions. (3) Confounder: Variables better handled through statistical adjustment rather than filtering. (4) Drop: Operational, subjective, redundant, or infeasible criteria in EHR data.

Formally, let *C* = {*c*_1_, *c*_2_, …, *c*_*n*_} denote the set of original eligibility criteria extracted from the clinical trial protocol. AERO learns a mapping *f*(*c*_*i*_, *K*) → *y*_*i*_, where *K* represents external clinical knowledge sources (e.g., UpToDate, pharmacological knowledge bases, and clinical guidelines), and *y*_*i*_ ∈ {Inclusion,Exclusion,Confounder,Drop} is the assigned category for criterion *c*_*i*_.

In practice, the model performs this classification by reasoning over a structured AI context that combines the candidate criterion with protocol intent and external clinical knowledge. For each eligibility criterion *c*_*i*_, the LLM constructs a reasoning context defined as Context_*i*_ = (*c*_*i*_, *P, K, O*), where *c*_*i*_ represents the candidate criterion extracted from the trial protocol, *P* denotes the protocol-level clinical intent, *K* represents external clinical knowledge retrieved from medical knowledge bases, and *O* represents operational feasibility considerations identified during LLM reasoning. This AI-driven contextual reasoning allows the system to distinguish between criteria that define the study population, criteria that ensure safety, and variables that should instead be handled through statistical adjustment, thereby improving the compatibility of trial protocols with real-world EHR data.

The optimized eligibility criteria set is then constructed as *C*^∗^ = *C*_incl_ ∪ *C*_excl_ ∪ *C*_conf_, where *C*_incl_ defines the minimal target population, *C*_excl_ enforces essential safety restrictions, and *C*_conf_ contains variables retained for downstream statistical adjustment.

During cohort construction, patients *p*_*j*_ from the EHR dataset are included if they satisfy all inclusion criteria and none of the exclusion criteria: *p*_*j*_ ∈ *S* if (∀*c*_*i*_ ∈ *C*_incl_, *c*_*i*_(*p*_j_) = 1) ∧ (∀*c*_*k*_ ∈ *C*_excl_, *c*_*k*_(*p*_j_) = 0), where *S* denotes the resulting emulated trial cohort. Each eligibility criterion *c*_*i*_(⋅) is represented as a binary indicator function applied to a patient record: 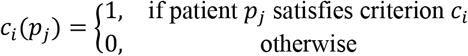. The first condition ∀*c*_*i*_ ∈ *C*_incl_, *c*_*i*_(*p*_*j*_) = 1 means that patient *p*_*j*_ must satisfy all inclusion criteria. For example, if the trial requires: age ≥ 18, diagnosis of heart failure, ejection fraction ≤ 35%, then the patient must meet every one of these conditions. The second condition ∀*c*_*k*_ ∈ *C*_excl_, *c*_*k*_(*p*_j_) = 0, means that patient *p*_*j*_ must not satisfy any exclusion criterion. Confounder variables *X* = {*x*_1_, *x*_2_, …, *x*_*m*_} identified by AERO are retained in the dataset and incorporated into downstream causal adjustment models (e.g., propensity score matching or Cox proportional hazards regression) rather than used as filtering criteria.

### Real-World Cohort Construction and Trial Emulation

The optimized criteria were applied to electronic health record data from the Mayo Clinic Platform (MCP), a large multi-institutional clinical data resource. Patients meeting the refined inclusion and exclusion criteria were selected to form treatment cohorts corresponding to the interventions studied in the original trial. Confounder variables identified by AERO were retained for downstream adjustment rather than cohort filtering. Trial emulation followed target trial principles, including definition of treatment groups, baseline time, follow-up, and outcome ascertainment. Only patients in the pre-trial completion period were included to avoid post-publication practice changes.

### Outcome Analysis

Comparative effectiveness between treatment groups was evaluated using a Cox proportional hazards model. The primary effect measure was the hazard ratio (HR) with corresponding p-value. Using the optimized criteria, the emulation produced an estimated HR of 1.561 (p = 0.0605). This result is consistent with the original trial findings, which also showed no statistically significant difference between treatments, indicating that AERO preserves the original trial conclusion while enabling real-world applicability.

## Results

### Study Details

To align with the design of the WARCEF trial, which was completed in July 2014, we restricted our real-world analysis to patients observed prior to 2014 in order to minimize potential changes in treatment patterns following trial dissemination. Study summaries and baseline characteristics for the MCP cohort before July 2014, the MCP cohort constructed using AERO-optimized criteria, and participants from the original WARCEF trial are presented in Figure 2. As shown in Panel A, the distribution of treatment assignment remains similar between the two cohorts, with aspirin representing the majority of patients in both groups, while the proportion of warfarin users remains relatively small. Gender composition differs modestly, with the adaptive cohort containing a lower proportion of male patients and a corresponding increase in female representation compared with the original cohort. Panel B shows that the adaptive cohort is older on average than the original cohort, with a higher mean age and comparable variability. Overall, these comparisons illustrate how the AERO-generated adaptive criteria produce a cohort that differs in certain demographic characteristics while preserving key treatment distributions, providing context for evaluating the relationship between randomized trial populations and real-world cohorts constructed through eligibility criteria optimization.

**Figure 2.**
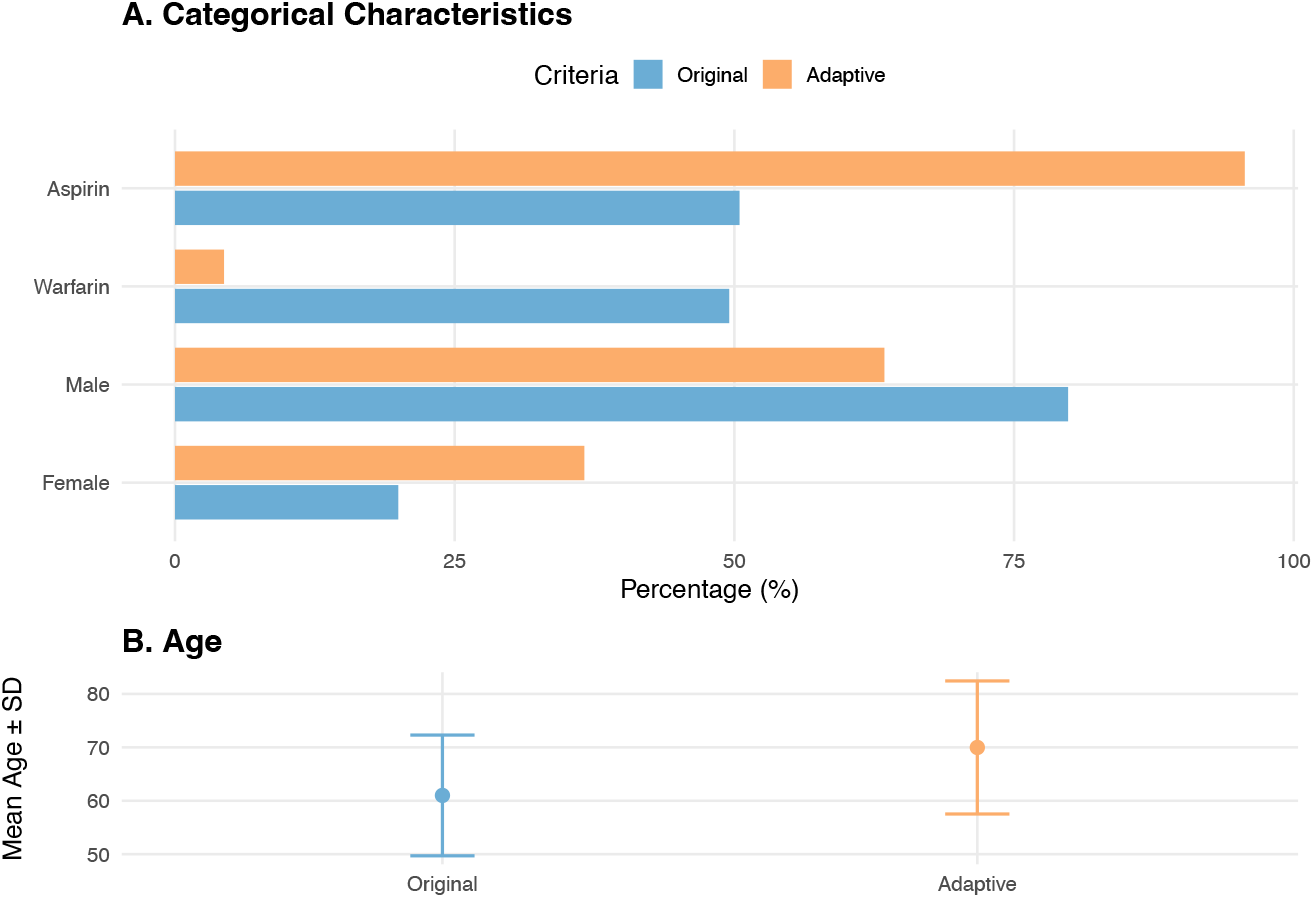
Cohort Characteristics Under Original and Adaptive Eligibility Criteria. Bar plots compare key demographic characteristics of cohorts constructed using the original trial eligibility criteria and the adaptive criteria generated by AERO. Panel A presents the percentage distribution of medication assignment (aspirin vs warfarin) and gender across the two cohorts. Percentages represent the proportion of patients in each cohort who fall into the corresponding category, calculated relative to the total number of patients in that cohort. Panel B shows the mean age with standard deviation for each cohort.

### Adaptive Categorization of Trial Eligibility Criteria

Table 2 summarizes how eligibility criteria from the original WARCEF trial protocol are categorized by the AERO framework for real-world trial emulation. Rather than treating all protocol conditions as strict filters, AERO organizes criteria into four functional categories: Strict Inclusion, Strict Exclusion, Confounder, and Drop / Operational. This structure distinguishes essential population-defining requirements from variables that should instead be handled analytically or removed due to lack of measurability in electronic health record (EHR) data.

**Table 2.**
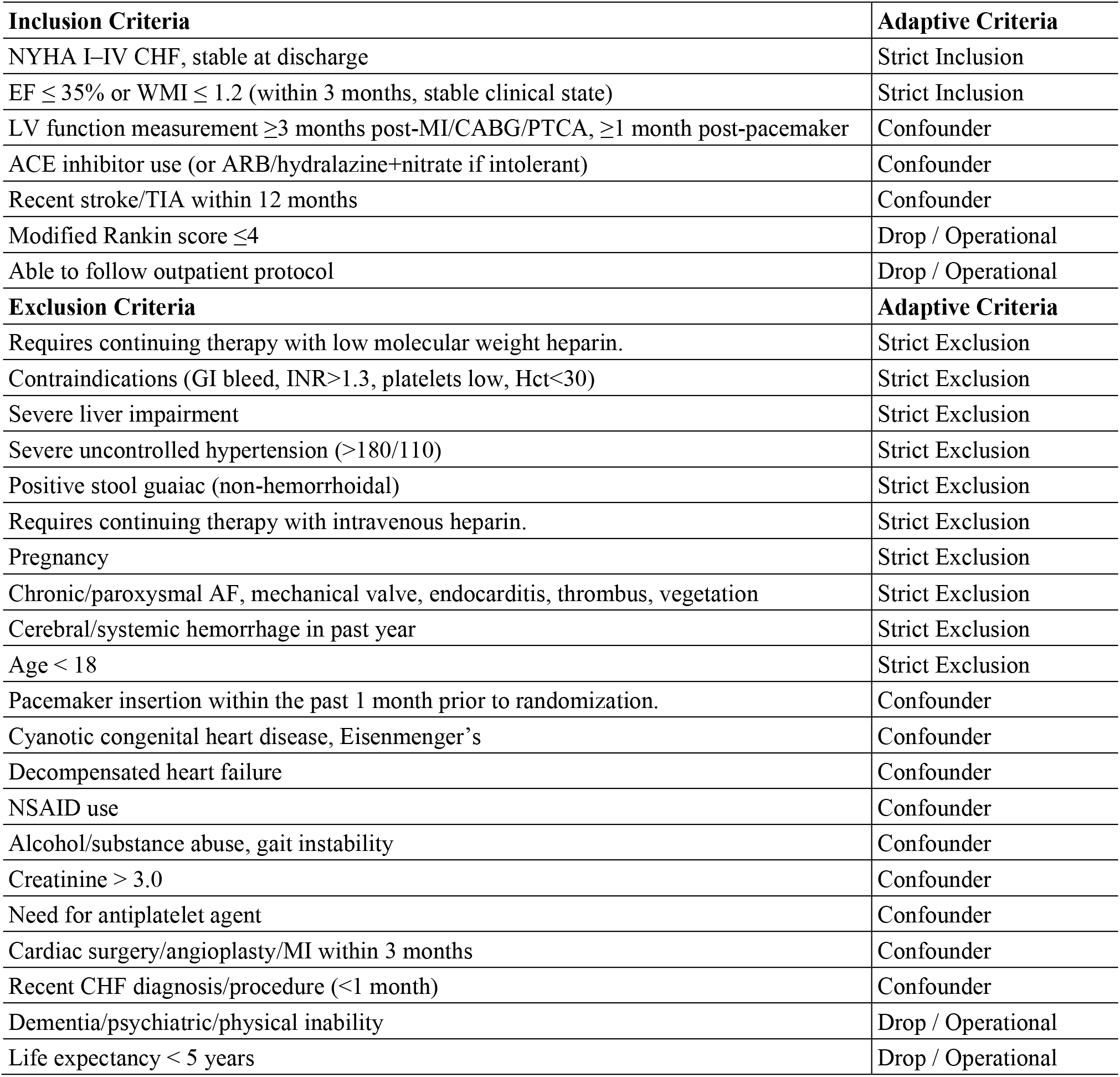
Adaptive Classification of Original Eligibility Criteria Using the AERO Framework. Eligibility criteria from the original WARCEF trial protocol are categorized by the AERO framework. Each criterion is assigned to one of four categories: Strict Inclusion, Strict Exclusion, Confounder, or Drop / Operational, based on clinical relevance, feasibility of extraction from electronic health record data, and potential influence on selection bias.

Core disease-defining conditions are retained as strict inclusion criteria, ensuring that the emulated cohort remains clinically consistent with the target trial population. In the WARCEF protocol, criteria related to heart failure diagnosis and left ventricular dysfunction, such as NYHA functional class and reduced ejection fraction, are preserved as strict inclusion requirements. These criteria define the primary disease population targeted by the original randomized trial.

Several clinically important contraindications are categorized as strict exclusions, reflecting their clear safety implications and reliable measurability in structured clinical data. Examples include severe liver impairment, severe uncontrolled hypertension, pregnancy, and major bleeding risk conditions. These exclusions maintain the safety boundaries of the original trial while ensuring that the resulting cohort remains clinically appropriate. Many clinical conditions that could influence treatment outcomes but do not necessarily define eligibility are categorized as confounders. These variables, including recent stroke, pacemaker insertion, Eisenmenger syndrome, decompensated heart failure, NSAID use, creatinine abnormalities, and need for antiplatelet therapy, are retained for statistical adjustment rather than used to restrict cohort inclusion. Treating such factors as confounders helps preserve sample size while allowing downstream analyses to control for clinical heterogeneity between treatment groups.

Finally, several protocol requirements are classified as Drop / Operational because they cannot be reliably operationalized in retrospective EHR data or primarily reflect study logistics rather than patient characteristics. These include criteria related to functional assessments, patient ability to adhere to study procedures, or life-expectancy judgments that are not consistently captured in routine clinical records. Overall, the AERO framework translates the original trial eligibility criteria into a structured and operationalizable format for observational data analysis. By distinguishing strict eligibility conditions from confounding variables and non-measurable operational requirements, this categorization supports robust real-world trial emulation while preserving the clinical intent of the original study design.

### Impact of Eligibility Criteria Optimization on Survival Outcomes and Risk Structure

Figure 3 (a) presents the Kaplan–Meier survival curves comparing aspirin and warfarin in the cohort constructed using the AERO-optimized eligibility criteria. The survival trajectories of the two treatment groups gradually diverge over time, with the aspirin group showing consistently higher survival probability than the warfarin group during follow-up. However, when treatment effect is estimated using a multivariable Cox proportional hazards model adjusted for the propensity score and additional clinical covariates, the association does not reach conventional statistical significance (p = 0.0605). These results indicate that although differences in survival patterns are observable over time, the adjusted treatment effect remains statistically inconclusive after accounting for baseline patient characteristics. Importantly, this finding is consistent with the conclusions of the original WARCEF randomized controlled trial, which reported no significant difference in overall mortality between warfarin and aspirin among patients with reduced ejection fraction. Together, these results suggest that the cohort constructed using the AERO framework preserves the essential clinical characteristics of the target trial population while enabling evaluation of treatment effectiveness using real-world observational data.

**Figure 3.**
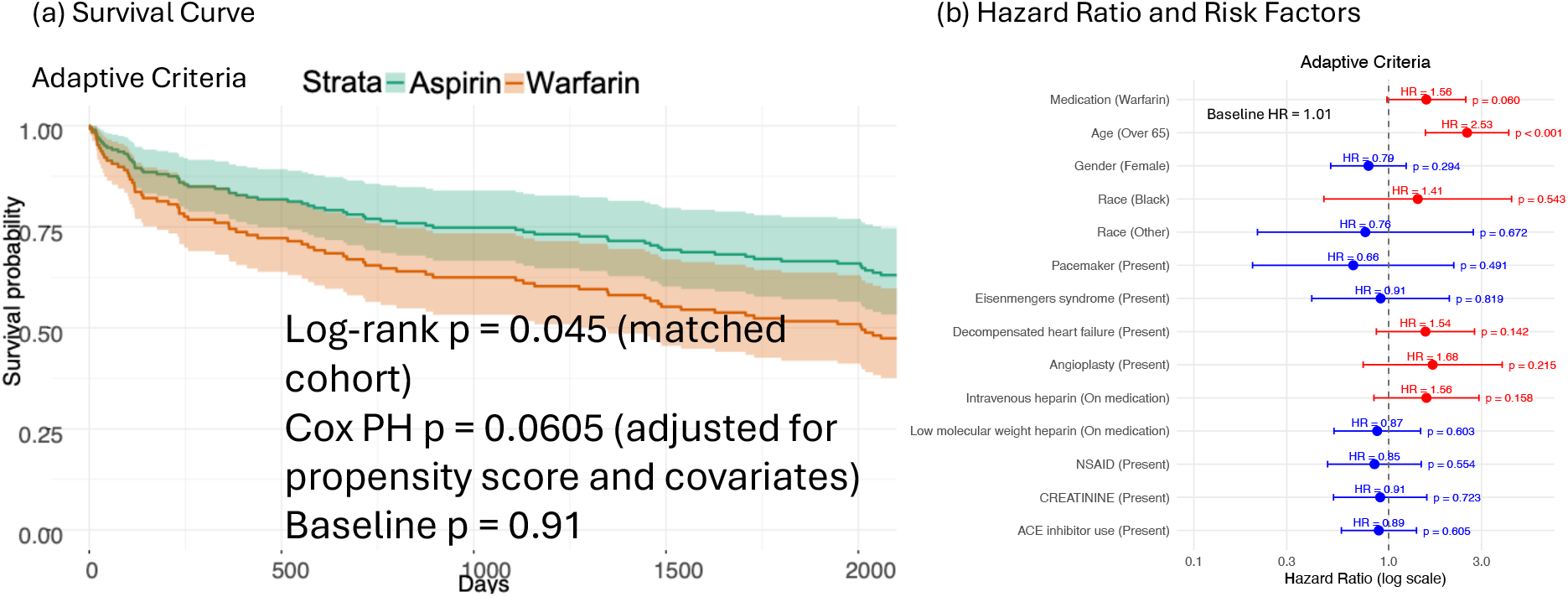
Kaplan–Meier Survival Curves Under Adaptive Eligibility Criteria and Hazard Ratios of Clinical Risk Factors in the Adaptive Eligibility Cohort. (a) Kaplan–Meier survival curves comparing aspirin and warfarin in the cohort constructed using AERO-optimized eligibility criteria from pre-2014 Mayo Clinic Platform data. The aspirin group demonstrates higher survival probability over time compared with the warfarin group. Shaded regions represent 95% confidence intervals. The treatment effect is estimated using a multivariable Cox proportional hazards model adjusted for the propensity score and additional covariates (p = 0.0605). The observed pattern remains consistent with the neutral findings reported in the original WARCEF randomized controlled trial. The log-rank p-value shown in each panel reflects the unadjusted comparison of survival distributions between treatment groups within the matched cohort. The Cox proportional hazards (PH) p-value represents the treatment effect estimated from a multivariable Cox model adjusted for the propensity score and additional covariates. Differences between the log-rank and Cox p-values may occur because the Cox model accounts for covariate adjustment, whereas the log-rank test provides an unadjusted comparison of survival curves. (b) Forest plot displaying hazard ratios (HRs) with 95% confidence intervals for mortality estimated from the Cox proportional hazards model in the cohort constructed using AERO-optimized eligibility criteria. Variables are shown on a logarithmic hazard ratio scale. Red markers indicate factors associated with increased mortality risk (HR > 1), whereas blue markers indicate protective associations (HR < 1).

Figure 3 (b) further illustrates the estimated hazard ratios for clinical variables included in the optimized eligibility cohort. Positive hazard ratios (red markers) indicate factors associated with increased mortality risk, whereas negative associations (blue markers) indicate protective effects. Age over 65 demonstrates the strongest association with increased mortality risk, followed by warfarin use, angioplasty history, intravenous heparin use, decompensated heart failure, and Black race. In contrast, several variables are associated with reduced mortality risk, including pacemaker presence, female sex, race categorized as Other, low molecular weight heparin use, ACE inhibitor use, and NSAID use.

Overall, the distribution of hazard ratios reflects the contribution of demographic characteristics, comorbid conditions, and medication use to survival outcomes within the optimized cohort. These results illustrate how the refined eligibility structure produced by AERO supports clinically interpretable risk modeling while maintaining a balanced cohort for real-world trial emulation.

### Ablation Analysis of Eligibility Handling Strategy

To ensure adequate sample size and statistical power, we treated the criteria “requires continuing therapy with low molecular weight heparin (LMWH),” “contraindications (e.g., gastrointestinal bleeding, INR > 1.3, low platelet count, hematocrit < 30),” and “requires continuing therapy with intravenous heparin” as confounders for adjustment rather than applying them as strict exclusion criteria. To further evaluate the impact of eligibility operationalization, we conducted an ablation analysis focusing on a representative criterion: the requirement for ongoing LMWH therapy. This criterion was examined under two alternative implementations: (a) modeled as a confounder, where LMWH use was retained in the cohort and adjusted for in the statistical model, and (b) applied as a strict exclusion criterion during cohort construction. Because excluding this criterion does not result in a drastic reduction in cohort size, it provides a practical example for evaluating how different eligibility handling strategies influence treatment effect estimates. In contrast, reclassifying several other exclusion criteria as confounders would lead to substantial reductions in the eligible population and make the resulting analyses infeasible. For this reason, LMWH requirement was selected as a representative case for the ablation study. We first modeled LMWH use as a confounder to preserve cohort size and population heterogeneity and then compared the resulting outcomes with those obtained when the same criterion was implemented as a strict exclusion.

As shown in Figure 5 (a), when the requirement for ongoing LMWH therapy is applied as a strict exclusion criterion, the Kaplan–Meier survival curves show a clearer separation between the warfarin and aspirin treatment arms (Cox proportional hazards p = 0.0347 after adjustment for propensity score and covariates). This apparent increase in treatment separation arises because excluding patients requiring LMWH removes a subgroup with more severe clinical conditions, thereby altering the baseline risk profile of the cohort. As a result, the observed difference between treatment groups may partly reflect changes in cohort composition rather than intrinsic differences in treatment efficacy. Figure 5 (b) presents the multivariable Cox proportional hazards model estimated under this strict exclusion setting, displaying the hazard ratios and confidence intervals for treatment and clinical covariates. The model indicates that multiple patient characteristics contribute to outcome risk alongside the treatment variable, highlighting the influence of underlying clinical factors on survival outcomes. Together, these results demonstrate that the way eligibility criteria are operationalized can materially influence comparative effectiveness estimates. In particular, applying clinically relevant but non-safety criteria as strict exclusions can introduce selection bias by disproportionately removing higher-risk patients and reshaping the study population. These findings emphasize the importance of systematic and bias-aware eligibility refinement when translating randomized trial protocols into real-world EHR data environments.

**Figure 5.**
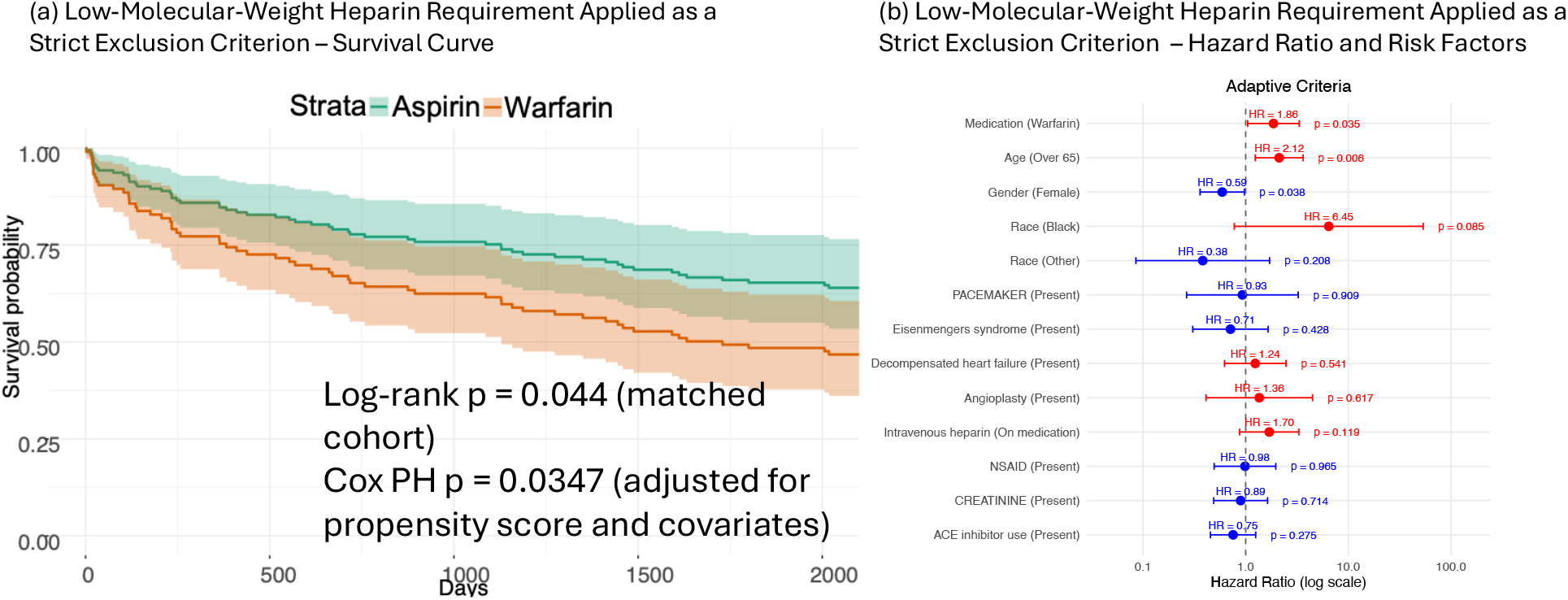
Ablation Study: Impact of Modeling Low-Molecular-Weight Heparin (LMWH) Requirement as a Confounder vs. Strict Exclusion. This figure illustrates how different operationalizations of the requirement for ongoing low-molecular-weight heparin (LMWH) therapy affect trial emulation results. Panel (a) shows Kaplan–Meier survival curves comparing aspirin and warfarin when LMWH use is treated as a strict exclusion criterion during cohort construction, resulting in a statistically significant separation between treatment groups (Cox PH p = 0.0347). Panel (b) presents the corresponding hazard ratio estimates and risk factors under the same strict exclusion setting. In contrast, when LMWH requirement is modeled as a confounder rather than a filtering rule, the survival difference between treatment arms is attenuated and becomes non-significant (p = 0.0605). These results demonstrate that the way eligibility criteria are operationalized, either as cohort exclusions or as variables adjusted during analysis, can substantially influence observed treatment effects. The ablation study highlights the importance of carefully distinguishing between exclusion criteria and confounders when adapting randomized trial protocols for real-world trial emulation. The log-rank p-value shown in each panel reflects the unadjusted comparison of survival distributions between treatment groups within the matched cohort. The Cox proportional hazards (PH) p-value represents the treatment effect estimated from a multivariable Cox model adjusted for the propensity score and additional covariates.

## Discussion

In this study, we developed and evaluated AERO, an AI agent–based framework designed to systematically refine clinical trial eligibility criteria for real-world trial emulation. While randomized controlled trials (RCTs) remain the gold standard for causal inference due to randomization and control of confounding ^1, 2^, restrictive eligibility criteria frequently limit generalizability to broader clinical populations ^20^. In particular, overly restrictive eligibility criteria have been shown to substantially reduce representativeness of trial participants compared with real-world populations ^21^. The increasing use of RWE derived from EHRs and large healthcare databases has expanded opportunities for evaluating treatment effectiveness in routine practice settings ^10^. Regulatory agencies have also emphasized the growing role of RWE in assessing drug safety and effectiveness ^7, 10^. However, robust RWE generation requires principled methodological alignment between trial design and observational implementation. Target trial emulation provides such a framework by explicitly specifying eligibility, treatment strategies, follow-up, and causal contrasts prior to analysis ^11, 13^. Yet, operationalizing eligibility criteria remains one of the most challenging aspects of emulation, particularly when criteria are partially unmeasurable or primarily logistical in nature ^22^.

Our findings demonstrate that eligibility operationalization is itself a design decision with causal implications. The ablation analysis showed that reclassifying ongoing low-molecular-weight heparin therapy from a confounder to a strict exclusion resulted in statistically significant treatment separation. This shift was driven not by mechanistic differences in treatment efficacy but by altered cohort composition. From a causal inference perspective, conditioning on restrictive eligibility criteria can reshape baseline risk distributions and induce bias through selection mechanisms ^23^. These results underscore that eligibility handling is not merely a preprocessing step but an integral component of causal study design ^24^.

AERO contributes to the methodological landscape in three primary ways. First, it formalizes eligibility adaptation as a structured reasoning task supported by heterogeneous clinical knowledge sources, rather than relying solely on manual heuristics. Second, it introduces a transparent taxonomy separating strict inclusion, safety exclusions, confounders, and operational artifacts, thereby enhancing reproducibility and interpretability. Third, it demonstrates that agentic AI systems can operate upstream of statistical modeling to improve study design without altering downstream analytical frameworks. In our case study, emulation using optimized criteria preserved the neutral treatment conclusion of the original WARCEF trial ^15^, supporting the validity of the approach while improving operational feasibility in real-world data.

Several limitations warrant discussion. First, the framework was evaluated using a single trial case study ^15^; broader validation across therapeutic areas and trial designs is necessary. Second, although AERO incorporates knowledge augmentation, it relies on large language model reasoning, which may be sensitive to prompt structure and knowledge retrieval completeness ^25^. Third, residual confounding and measurement error inherent in observational EHR data cannot be entirely eliminated, even under target trial emulation principles ^26^. Finally, eligibility reclassification decisions may require contextual clinical judgment and may not generalize uniformly across disease domains. Another limitation of the AERO framework is the potential introduction of selection bias during eligibility criteria optimization. By restructuring and relaxing certain trial criteria to improve feasibility within electronic health record data, the resulting cohort may differ from the population originally targeted in the randomized trial.

Despite these limitations, our results demonstrate that systematic, knowledge-informed eligibility refinement can preserve trial-level conclusions while improving real-world applicability. As the role of RWE continues to expand, scalable and transparent methods for eligibility translation will be critical for bridging the gap between controlled clinical trials and routine clinical care.

## Conclusion

AERO provides a systematic, AI-driven framework for translating randomized trial eligibility criteria into operational definitions suitable for real-world data. By distinguishing core population definitions, safety exclusions, confounders, and non-measurable operational artifacts, AERO reduces infeasible restrictions and mitigates selection bias while preserving clinical intent. Through application to the WARCEF trial, we demonstrate that eligibility optimization reshapes cohort composition and prognostic structure without altering the primary comparative conclusion. The ablation analysis further shows that eligibility handling decisions can materially influence observed treatment effects, emphasizing the importance of principled criteria adaptation in target trial emulation. Together, these findings position AERO as a scalable and reproducible methodology for AI-assisted evidence generation. As real-world data continues to expand in scope and regulatory relevance, agentic systems like AERO may play a critical role in bridging the gap between controlled clinical trials and routine clinical practice, supporting more transparent, adaptable, and clinically grounded causal inference.

## Data Availability

All data produced in the present study are not available publicly. They are Mayo's internal data.

